# PERMISSIVE OMICRON BREAKTHROUGH INFECTIONS IN INDIVIDUALS WITH BINDING OR NEUTRALIZING ANTIBODIES TO ANCESTRAL SARS-CoV-2

**DOI:** 10.1101/2022.04.17.22273938

**Authors:** Erin Williams, Jordan Colson, Ranjini Valiathan, Juan Manuel Carreño, Florian Krammer, Michael Hoffer, Suresh Pallikkuth, Savita Pahwa, David Andrews

## Abstract

**Background:** Breakthrough infection with the severe acute respiratory syndrome coronavirus 2 (SARS-CoV-2) Omicron variant (B.1.1.529) has occurred in populations with high vaccination rates. These infections are due to sequence variation in the spike protein leading to a reduction in protection afforded by the current vaccines, which are based on the original Wuhan-Hu-1 strain, or by natural infection with pre-Omicron strains.

**Methods:** In a longitudinal cohort study, pre-breakthrough infection sera for Omicron breakthroughs (n=12) were analyzed. Assays utilized include a laboratory-developed solid phase binding assay to recombinant spike protein, a commercial assay to the S1 domain of the spike protein calibrated to the World Health Organization (WHO) standard, and a commercial solid-phase surrogate neutralizing activity (SNA) assay. All assays employed spike protein preparations based on sequences from the Wuhan-Hu-1 strain. Participant demographics and clinical characteristics were captured.

**Results:** Pre-breakthrough binding antibody (bAB) titers ranged from 1:800-1:51,200 for the laboratory-developed binding assay, which correlated well and agreed quantitatively with the commercial spike S1 domain WHO calibrated assay. SNA was detected in 10/12 (83%) samples.

**Conclusions:** Neither high bAB nor SNA were markers of protection from Omicron infection/re-infection. Laboratory tests with antigen targets based on Wuhan-Hu-1 may not accurately reflect the degree of immune protection from variants with significant spike protein differences. Omicron breakthrough infections are likely due to high sequence variation of the spike protein and reflect incomplete immune protection from previous infection with strains that preceded Omicron or with vaccinations based on the original Wuhan-Hu-1 strain.

## INTRODUCTION

First reported in November 2021, the Omicron (B.1.1.529/BA.1) variant of SARS-CoV-2 progressed rapidly to become the predominant strain in the United States, comprising 99.5% of all new infections [1] in mid-December 2021. The Omicron variant is notable for its high transmissibility within the population with pre-existing immunity, as well as significant antigenic differences in the spike protein compared to the original Wuhan-Hu-1 strain as well as variants like Alpha, Beta, Gamma and Delta [2]. The Omicron spike (S) glycoprotein, which is responsible for initiating viral entry into cells [3], contains at least 32 amino acid changes [4] compared to the Wuhan-Hu-1 strain, with mutations spanning the receptor binding domain (RBD) and N-terminal domain (NTD) and the furin cleavage site (FCS).

Most severe acute respiratory syndrome coronavirus 2 (SARS-CoV-2) vaccines currently available are based on spike protein sequences derived from the Wuhan-Hu-1 strain. Consequently, as variant SARS-CoV-2 strains have emerged with altered spike protein sequences (compared to the Wuhan-Hu-1 strain), reduced antibody binding titers and reduced viral neutralizing activities have been observed [5]. Recent work has shown that a reduction in antibodies with spike binding and viral neutralizing activity [6-10] are associated with an increased risk of infection in fully vaccinated individuals, including those who were previously infected.

Along with nucleic acid and antigen testing, antibody testing has been widely utilized in research, epidemiological and clinical settings during the coronavirus disease 2019 (COVID-19) pandemic, with over 85 tests commercially available under US FDA Emergency Use Authorization to date [11]. The majority of the antibody detection methods employ solid phase binding assays such as enzyme linked immunosorbent assays (ELISA), with some capable of detecting surrogate neutralizing antibody activity [12]. Unfortunately, the role of antibodies as correlates of protection, while well established in general [13-16], has been complicated by the lack of standardization for US serology assays and the emergence of variants [17].

Antibody reactivity to recombinant spike proteins has been widely used as a marker for humoral immunity [16, 18-20], particularly in the context of the recent Delta and Omicron waves, both associated with breakthrough infections worldwide. Several groups have reported the use of binding or viral neutralizing antibody titers as correlates of protection against SARS-CoV-2 [16]. Similarly, there is evidence to support an association between lower viral neutralizing antibody titers and breakthrough infections [17, 21, 22], though a specific threshold for risk reduction or protection has not been well established. Increased viral neutralizing activity has also been observed following booster vaccination [23, 24], with associated reduced disease severity and rates of hospitalization, in comparison with individuals who did not receive booster doses.

Here, we investigate binding antibody titers and receptor binding domain (RBD)-angiotensin converting enzyme 2 (ACE2) interaction inhibiting antibody activity among previously vaccinated patients presenting with presumptive Omicron breakthrough infection. Our study compares three antibody tests: a two-step quantitative IgG binding assay to the full spike ectodomain (Icahn School of Medicine at Mount Sinai assay), a semi-quantitative assay for total serum immunoglobulins inhibiting RBD-ACE2 interactions (GenScript cPass)[25], and a quantitative binding titer assay for IgG to S1 domain (Ortho Clinical Diagnostics VITROS)[26], which is the only commercial antibody assay calibrated to the WHO standard in the United States. The primary aim of this work is to investigate the impact of binding antibody titers and RBD-ACE2 interaction inhibition titers on Omicron breakthrough infections. We also aim to assess the limitations of the three clinically available laboratory tools in the context of Omicron variant infection, which is known to have significant properties of immune escape.

## METHODS

### Study Design and Participants

We included 12 study participants who are enrolled in our IRB-approved (#20201026), ongoing, longitudinal SARS-CoV-2 immunity study (“CITY”) at the University of Miami Miller School of Medicine. Following written informed consent, participants answered a demographic and health history questionnaire. Nasal swab samples (Ruhof, Mineola, NY) were collected at each visit to screen for active SARS-CoV-2 infection and whole blood samples (Becton Dickinson, Franklin Lakes, NJ) were drawn for serum before storage at -80°C. All participants agreed to sample banking and consented to their use in future research. Any individual (12/186 active participants [6.5%]) who self-reported breakthrough infection between December 15^th^, 2021, and January 7^th^, 2022, was included in this study. They were polled for additional information regarding their breakthrough infection, including associated symptoms.

### Omicron Breakthroughs

Previously vaccinated individuals who experienced breakthrough infection between December 15^th^, 2021, and January 7^th^, 2022, were included in this study (n=12). These dates corresponded with the national surge associated with high (>98%) Omicron variant infection prevalence. Breakthrough infections were established with a clinically validated positive nucleic acid amplification test (NAAT) method, such as PCR (polymerase chain reaction). Serum samples obtained at the study visit prior to breakthrough infection were retrieved from storage for the individuals described above.

### Assays

#### Mount Sinai Laboratory assay

The SARS-CoV-2 ELISAs were performed using a well-described two step assay developed by the Icahn School of Medicine at Mount Sinai [27-29]. Briefly, 96-well plates were coated at 4°C with SARS-CoV-2 spike protein (2 µg/ml) solution and incubated overnight. Plates were blocked with 3% non-fat milk prepared in PBS with 0.1% Tween 20 (PBST) and incubated at room temperature for 1h. After blocking, serial dilutions of heat inactivated serum samples were added to the plates and incubated for 2h at room temperature. Plates were washed three times with 0.1% PBST followed by addition of a 1:3,000 dilution of goat anti-human IgG–horseradish peroxidase (HRP) conjugated secondary antibody (50μl) well and incubated 1h. Plates were washed, 100 µl SIGMAFAST OPD (o-phenylenediamine dihydrochloride;) solution was added to each well for 10 min and then the reaction was stopped by the addition of 50μl per well of 3M hydrochloric acid. The optical density at 490nm (OD490) was measured using a Synergy 4 (BioTek) plate reader. The background value was set at an OD490 of 0.15 then discrete titers were reported in values of 1:100, 1:200, 1:400, 1:800, 1:1600, 1:3200, 1:6400, 1:12800, 1:25600, 1:51200, 1:102400, and 1:204800. The limit of detection was set at 1:100.

#### Genscript cPass™ surrogate neutralization antibody assay

This semi-quantitative SARS-CoV-2 surrogate neutralizing antibody assay, which measures the inhibition of RBD and ACE2 interactions, was performed in accordance with manufacturer’s instructions. In brief, participant serum was mixed with dilution buffer and soluble RBD-HRP conjugate. After a 30-minute incubation at 37 °C, samples were added to a 96-well plate which was pre-coated with recombinant human ACE2 protein. The plate was incubated for 15 min at 37 °C, sample mixture removed, wells were washed, substrate added, and plates were read at 450nm on a Dynex (Dynex Technologies, Chantilly, VA) Agility multiplate ELISA instrument. Data was reported as percent neutralization with a threshold of 30% as the cutoff for surrogate neutralizing activity. Values from 30-100% surrogate neutralizing activity were considered positive and values below 30% were considered negative for surrogate neutralizing activity.

#### Ortho Clinical VITROS SARS-CoV-2 IgG assay

This assay was performed following the manufacturer’s instructions on an Ortho-Clinical VITROS 7600 analyzer (Ortho Clinical Diagnostics, Raritan, NJ). This assay is calibrated to the 1^st^ WHO International Standard Anti-SARS-CoV02 Immunoglobulin (Human), NIBSC [26] with results reported in the WHO standard units of IgG Binding Antibody Units/ml (BAU/ml) to recombinant spike S1 domain. The assay measurement range is 2.0 – 200 BAU/ml. Participant serum was diluted with manufacturer diluent to achieve a measurable result within the manufacturer measurement range, followed by conversion of the result with the dilution factor to achieve the final BAU/ml concentration.

### Statistical analysis

Following log-transformation, Pearson correlation coefficients and Bland-Altman plots were generated in order to examine the correlation and degree of agreement between the Mount Sinai Laboratory assay and Ortho-Clinical VITROS assay. All statistical analyses and figures generated were performed in R Studio.

## RESULTS

### Patient characteristics

As shown in Table 2, the gender distribution was equal with a median age of 50.5 (range: 30-78). Six (50%) entered the study with a previous history of COVID-19, confirmed via documented NAAT testing. The participants most commonly identified as White (83.3%) and non-Hispanic (66.6%). None were known to be immunocompromised. All had received primary vaccination greater than 90 days prior to breakthrough, to include Pfizer/BioNTech BNT162b2 (66.7%), Moderna mRNA-1273(25%), and Johnson & Johnson Ad26.COV2.S (8.3%). Among those patients who had not yet received a booster (n=5), two reported a history of previous COVID-19 at study entry. Also of note, the one individual who received the Johnson & Johnson vaccine had been previously infected with SARS-CoV-2 twice prior to vaccine receipt. Seven received a booster vaccine (58.3%), with 2 receiving Pfizer/BioNTech BNT162b2 (28.6%) and 5 receiving Moderna mRNA-1273 (71.4%). Three of the participants who opted for a Moderna mRNA-1273 booster received a half dose (i.e., 50 mcg vs. 100 mcg).

**Table 1:** Selected characteristics of the SARS-CoV-2 serological assays per laboratory or manufacturer specifications.

|  | **Mount Sinai 2-Step ELISA [27, 28, 30]** | **GenScript cPass [25]** | **Ortho-Clinical VITROS [26, 31]** |
| --- | --- | --- | --- |
| **Assay type** | ELISA/binding | ELISA/blocking | Solid phase/binding |
| **Antigen(s) used** | RBD + recombinant full-length spike | RBD | Spike S1 domain |
| **Antibody class detected** | IgG | Total (IgG/M/A) | IgG |
| **Units of detection** | Discrete dilution titers or continuous area under the curve values<sup>b</sup> | Percent surrogate neutralization activity (<30%, 30-100%)* | Binding antibody units/ml (BAU/ml) |
| **Specificity (95% CI)** | 100%<sup>a</sup> | 100%<sup>a</sup> | 100% (99.3-100.0%) |

RBD = receptor binding domain
<sup>a</sup> CI not provided
<sup>b</sup> AUC values were not included in this analysis

**Table 2:**
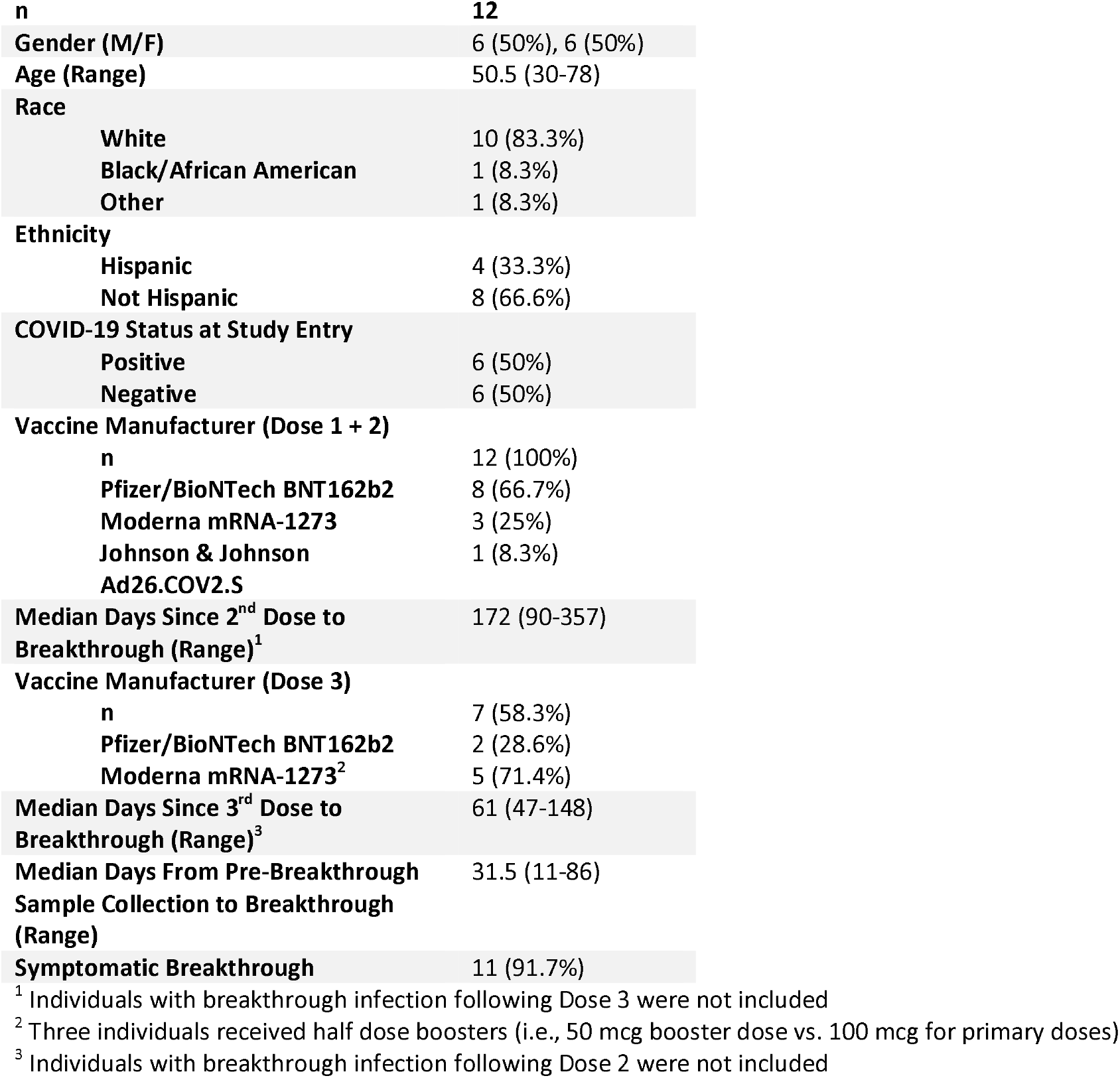
Participant characteristics for Omicron breakthrough infections.

### Breakthrough Infection Characteristics

All breakthroughs described above were classified as mild, given the limited symptoms and lack of medical intervention or hospitalization. As seen in Table 3, the most frequently reported symptoms were cough (66.6%) and congestion/rhinorrhea (66.6%). Sore throat (41.6%) and headache (41.6%) were also highly reported. Only one participant reported no symptoms. No participant reported nausea, vomiting, or dyspnea.

**Table 3:** Symptoms Reported During Omicron Breakthrough Infection.

| Participant | Cough | Fever | Congestion<br>Runny Nose | Sore Throat | Dyspnea | Anosmia | Dysgeusia | Upset Stomach<br>Diarrhea | Nausea<br>Vomiting | Fatigue | Myalgias | Headache |
| --- | --- | --- | --- | --- | --- | --- | --- | --- | --- | --- | --- | --- |
| 1<sup>a</sup> |  |  |  |  |  |  |  |  |  |  |  |  |
| 2 |  | X |  | X |  | X | X |  |  |  | X |  |
| 3 | X |  | X | X |  | X | X |  |  |  |  |  |
| 4 | X |  | X |  |  |  |  | X |  |  | X | X |
| 5<sup>b</sup> |  | X | X |  |  |  |  |  |  | X | X | X |
| 6<sup>b</sup> | X | X |  | X |  |  |  |  |  |  |  |  |
| 7<sup>b</sup> | X |  | X |  |  |  |  |  |  |  |  |  |
| 8<sup>b</sup> | X |  | X |  |  |  |  | X |  | X |  |  |
| 9<sup>b</sup> | X | X |  |  |  |  |  |  |  |  |  |  |
| 10 | X |  | X | X |  |  |  |  |  |  |  | X |
| 11<sup>b</sup> |  |  | X | X |  |  |  |  |  | X | X | X |
| 12<sup>b</sup> | X |  | X |  |  |  |  |  |  | X |  | X |
| **n=12** | **8 (66.6%)** | **4 (33.3%)** | **8 (66.6%)** | **5 (41.6%)** | **0 (0%)** | **2 (16.6%)** | **2 (16.6%)** | **2 (16.6%)** | **0 (0%)** | **4 (33.3%)** | **4 (33.3%)** | **5 (41.6%)** |

<sup>a</sup> Asymptomatic; no symptoms reported and was found to be positive after positive exposure
<sup>b</sup> Received a booster vaccination

### Assay Results

As shown in Table 4, all participants had detectable antibodies (discrete titers) with the Mount Sinai assay all had detectable antibodies (BAU/ml) with the VITROS assay (Figure 1). Titers ranged from 1:800 to 1:51,200 for the Mt. Sinai assay (median: 1:19200; range: 1:800-51200) and from 57.4 to 13,500 BAU/ml for the VITROS assay (median: 2710; range: 103 – 13500). The cPass RBD-ACE2 interaction inhibition assay showed 10/12 (83.3%) participants had positive surrogate neutralizing activity (SNA) with 2/12 (16.7%) lacking SNA (<30%) as determined by manufacturer specifications. Among those participants with positive surrogate neutralizing activity, 8/10 had 98% SNA, and two individuals had 90% SNA and 58% SNA respectively. Notably, the two individuals found negative for SNA were also found to have lower values for the binding assays (i.e., Mount Sinai and VITROS).

**Table 4:** Mt. Sinai Laboratory, Ortho-Clinical VITROS, and Genscript cPASS Assay Results.

| Participant | # of SARS-CoV-2 Challenges (Infection(s) and Vaccines) Prior to BT |  | Days Between Pre-Breakthrough Sample and Last Positive NAAT Test | Icahn SOM at Mt. Sinai Assay<sup>a</sup> | cPass Assay<sup>b</sup> | Ortho-Clinical VITROS<sup>c</sup> |
| --- | --- | --- | --- | --- | --- | --- |
| 1 | 3 | NI, V2 | 75 | 51200 | 98, POS | 6600 |
| 2 | 2 | UI, V2 | 33 | 800 | <30, NEG | 57.4 |
| 3 | 2 | UI, V2 | 16 | 1600 | 58, POS | 103 |
| 4<sup>f</sup> | 3 | NII, V1 | 11 | 1600 | 90, POS | 116 |
| 5<sup>d</sup> | 3 | UI, V3 | 27 | 51200 | 98, POS | 6160 |
| 6<sup>d</sup> | 3 | UI, V3 | 70 | 51200 | 98, POS | 7490 |
| 7<sup>d,e</sup> | 4 | NI, V3 | 23 | 12800 | 98, POS | 2250 |
| 8<sup>d,e</sup> | 4 | NI, V3 | 36 | 25600 | 98, POS | 2710 |
| 9<sup>d</sup> | 3 | UI, V3 | 86 | 51200 | 98, POS | 13500 |
| 10 | 3 | NI, V2 | 33 | 6400 | <30, NEG | 635 |
| 11<sup>d,e</sup> | 4 | NI, V3 | 30 | 25600 | 98, POS | 4890 |
| 12<sup>d</sup> | 3 | UI, V3 | 19 | 6400 | 98, POS | 1210 |

UI = No hx of natural infection prior to BT; NI = Hx of natural infection prior to BT; NII: Hx of natural infection 2x prior to BT; V1 = Vaccinated once; V2 = Vaccinated twice; V3 = Vaccinated twice + booster vaccination; BT = Breakthrough infection
<sup>a</sup> Discrete antibody titers may be reported from 1:100-204,800, though in this study discrete titers were only observed up to 51,200.
<sup>b</sup> Surrogate neutralizing activity was reported from <30 – 100% (< 30% = Neg, 30 – 100% = Pos)
<sup>c</sup> Undiluted linear range 2 – 200 BAU/ml, samples diluted as needed to achieve result in linear range of 2-200 BAU/ml
<sup>d</sup> Received a booster vaccination
<sup>e</sup> Received a Moderna mRNA-1273half dose
<sup>f</sup> Received Johnson & Johnson Ad26.COV2.S as primary vaccination

**Figure 1:**
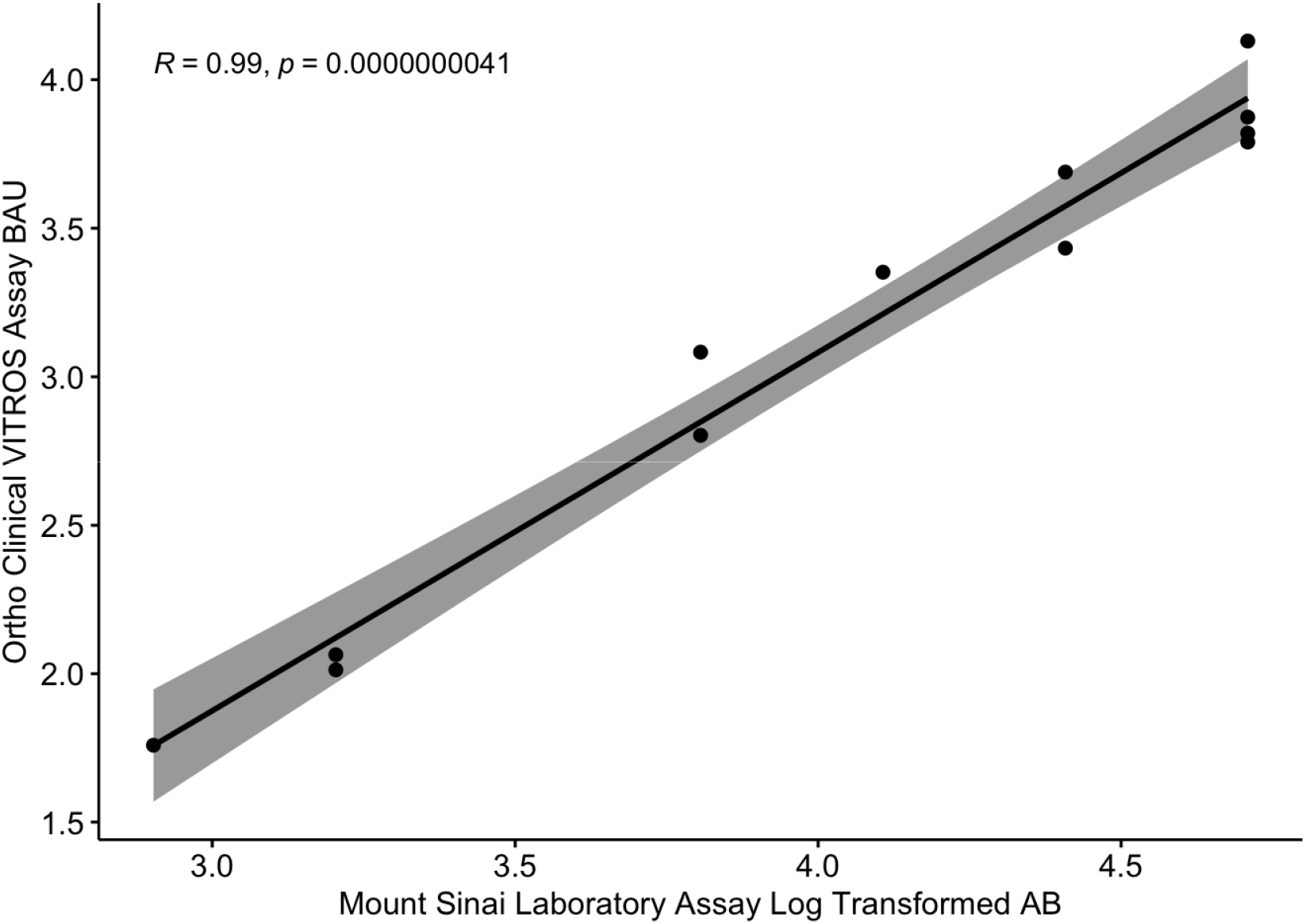
The Mount Sinai Laboratory Assay and the Ortho Clinical VITROS Assay are strongly correlated. *Discrete antibody titers from the Mount Sinai Laboratory assay and binding antibody units (BAU/ml) from the Ortho Clinical VITROS assay were log-transformed prior to analysis. The assays were strongly correlated (t* = 19, Pearson’s *r* = 0.99, 95% CI = 0.949 – 0.996; p = <0.001). The grey shaded area indicates the standard error margins.

### The Icahn School of Medicine at Mount Sinai and the Ortho-Clinical VITROS Assay Correlate Well and Quantitatively Agree

The participants’ log-transformed, pre-breakthrough antibody titers ranged from 2.90 to 4.71. Bland-Altman analysis demonstrated good agreement between assays, as shown in Figure 2. The mean difference was 0.91, with a 95% confidence interval of 0.527 to 1.285. Additionally, we found that the Mount Sinai Laboratory assay endpoints were strongly correlated with the Ortho-Clinical VITROS assay endpoints (Pearson correlation of r(10) = .99, p = <0.00001). We also examined the correlation between the Mount Sinai Laboratory assay and the cPass assay, and found that the two assays were positively associated (Pearson correlation of r(10) = .69, p = 0.013), though this was to a lesser degree than the VITROS assay.

**Figure 2:**
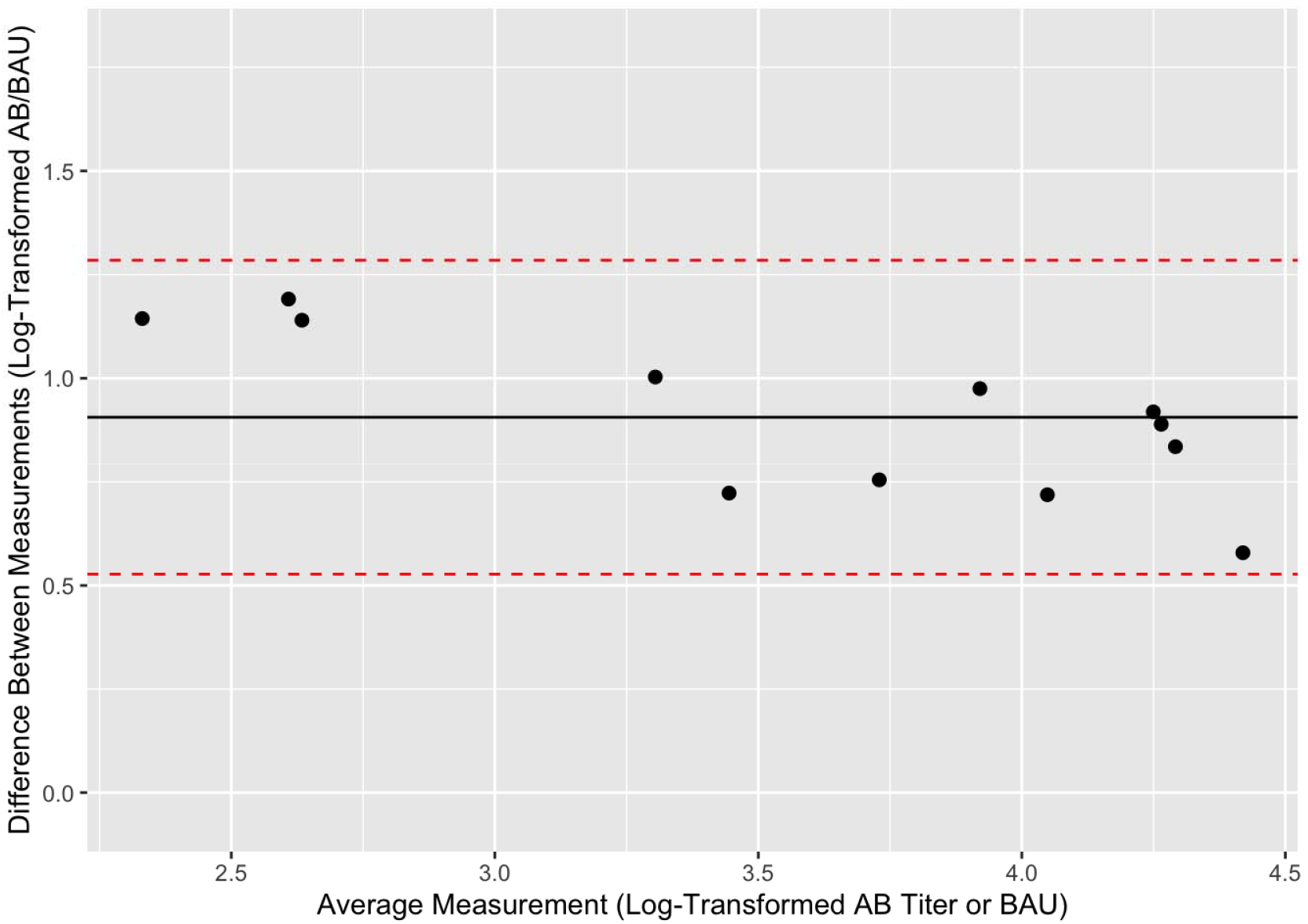
The Mount Sinai Laboratory Assay and the Ortho Clinical VITROS Assay quantitatively agree. A Bland-Altman plot was generated in order to describe agreement between the Mount Sinai Laboratory assay and the Ortho Clinical VITROS assays. The y-axis demonstrates the difference between paired, log-transformed measurements (i.e., discrete AB values from the Mount Sinai vs. BAU from the Ortho Clinical VITROS assay) for each participant. The x-axis represents the average of the log-transformed measurement from each assay. The mean difference (0.91) between values is indicated by the black line, while the red dotted line represents the 95% confidence interval limits (0.527 – 1.285) for the average difference between assays. All data points fell within the limits of agreement, indicating good agreement between the assays.

We designated the participants based on the number of SARS-CoV-2 “antigenic challenges” they have experienced (i.e., vaccination and/or natural infection). After review of the variation in combinations between natural infection and vaccination, individuals with only two SARS-CoV-2 “antigenic challenges” appeared to have lower antibody titers (or less reactivity) across all three assays, although there is no clear relationship between the quantity of “antigenic challenges” and increased antibody titers. Those who had received booster vaccinations had higher antibody titers than those who did not with the exception of one participant who had received their 2nd dose within 90 days of the breakthrough infection.

## DISCUSSION

The goal of this study was to investigate the impact of binding antibody titers and RBD-ACE2 interaction inhibition titers on Omicron breakthrough infections. The binding antibody assays (Mount Sinai Laboratory and Ortho-Clinical VITROS) correlated and quantitatively agreed. Given that the Ortho-Clinical VITROS assay is calibrated to the WHO standard, this finding adds additional validity to this and future work, particularly when conducting comparisons across assays. We also found that higher binding titers were generally suggestive of higher SNA, though unfortunately magnitude (percent neutralization) alone was clearly an insufficient marker of protection in preventing breakthrough infection for the cohort described above. Overall, our results indicate that the observation of high binding titers or SNA to ancestral spike/RBD alone do not adequately confer protection from breakthrough infection with the Omicron variant. In spite of a recent study[32] suggesting adequate viral neutralizing activity following booster vaccination, this study provides compelling evidence in a longitudinal cohort that despite robust detection of antibody levels against the ancestral-strain by three distinct assays, this does not establish proof of sufficient protection against antigenically distant variants.

In this study, we have compared three distinct antibody assays in a vaccinated and/or boosted population of suspected Omicron breakthrough cases. This work has several limitations, principally the small sample size and variable immune experience of the cohort. Further, as is the case for the majority of the available antibody assays, the Mount Sinai Laboratory assay and the cPass assay have not yet been calibrated to the WHO standard. Our study does provide evidence for a correlation between the Mount Sinai Laboratory and Ortho-Clinical VITROS assays, however. We believe that further standardization of the serological assays to an international standard will allow better correlations of immunity between independent clinical trials.

Finally, this study examined only samples immediately prior to breakthrough infection so factors regarding temporal relationship of infection, clinical presentation, and sample collection may have affected our observations. Additional studies including individuals who appear to be susceptible to re-infection or who are poor immunologic responders to SARS-CoV-2 infection and/or vaccination, are needed to better understand differential immune kinetics in those populations. Of particular interest will be characterizing the T-cell immune responses to SARS-CoV-2 in individuals who have developed strong antibody responses yet experience breakthrough infection. Importantly, since we did not compare the titers of breakthrough cases with titers of non-breakthrough cases, we cannot draw conclusions regarding where these titers would fall and if they trend lower than those of individuals who may have been exposed but were not infected.

Although mounting evidence suggests that both primary vaccination and boosters lessen the likelihood of symptomatic infection, hospitalization and death following infection with Omicron, there remains an urgent need for updated variants-specific an ultimately “variant-proof” vaccines and early treatment modalities. This work has also highlighted the need for large-scale harmonization across serological assays, particularly those that have low barriers for use (e.g., low cost, time-efficient), and that can be scaled expediently to the population level. Further, COVID-19 severity, particularly in the case of breakthrough infections, is related to antigen-specific adaptive immune responses [33-36], though the precise mechanisms for individual susceptibility to breakthrough infections remain unclear. In addition to greater clarification on the specifics of the humoral response to infection and vaccination is needed – especially the role of mucosal antibodies and non-neutralizing antibodies. Future work should incorporate profiling of cellular immunity to better define the immune landscape amongst diverse populations affected by SARS-CoV-2 and its variants.

## Data Availability

All data produced in the present work are contained in the manuscript.

## Funding

This work was partly funded by the NIAID Collaborative Influenza Vaccine Innovation Centers (CIVIC) contract 75N93019C00051 as part of the PARIS/SPARTA studies.

## Acknowledgements

As ever, we are grateful to our research participants for their samples, time, and interest in this work. At the University of Miami Miller School of Medicine, we thank Margaret Roach, Elizabeth Varghese, and the entire phlebotomy team at the Clinical Translational Research Site. We would also like to thank the SARS-CoV-2 serology team at the Icahn School of Medicine at Mount Sinai, including Dominika Bielak and Gagandeep Singh.

## Conflicts of Interest

The Icahn School of Medicine at Mount Sinai has filed patent applications relating to SARS-CoV-2 serological assays (U.S. Provisional Application Numbers: 62/994,252, 63/018,457, 63/020,503 and 63/024,436) and NDV-based SARS-CoV-2 vaccines (U.S. Provisional Application Number: 63/251,020) which list Florian Krammer as co-inventor. Patent applications were submitted by the Icahn School of Medicine at Mount Sinai. Mount Sinai has spun out a company, Kantaro, to market serological tests for SARS-CoV-2. Florian Krammer has consulted for Merck and Pfizer (before 2020), and is currently consulting for Pfizer, Third Rock Ventures, Merck, Seqirus and Avimex. The Krammer laboratory is also collaborating with Pfizer on animal models of SARS-CoV-2.

All other authors declare that they have no known competing financial interests or personal relationships that could have appeared to influence the work reported in this paper.

## Notes

### Author Declarations

IRB of University of Miami Miller School of Medicine gave ethical approval for this work (#20201026).

